# SARS-CoV-2 Testing in Florida, Illinois, and Maryland: Access and Barriers

**DOI:** 10.1101/2020.12.23.20248789

**Authors:** Steven J. Clipman, Amy Wesolowski, Shruti H. Mehta, Smisha Agarwal, Sarah E. Cobey, Derek A.T. Cummings, Dustin G. Gibson, Alain B. Labrique, Gregory D. Kirk, Sunil S. Solomon

**Affiliations:** Department of International Health, Johns Hopkins Bloomberg School of Public Health, Baltimore, Maryland, United States of America; Department of Epidemiology, Johns Hopkins Bloomberg School of Public Health, Baltimore, Maryland, United States of America; Department of Ecology and Evolution, University of Chicago, Chicago, Illinois, United States of America; Department of Biology and the Emerging Pathogens Institute, University of Florida, Gainesville, Florida, United States of America; Department of Medicine, Division of Infectious Diseases, Johns Hopkins University School of Medicine, Baltimore, Maryland, United States of America

## Abstract

**Objective:** To characterize the SARS-CoV-2 testing cascade and associated barriers in three US states.

**Methods:** We recruited participants from Florida, Illinois, and Maryland (∼1000/state) for an online survey September 16 – October 15, 2020. The survey covered demographics, COVID-19 symptoms, and experiences around SARS-CoV-2 PCR testing in the prior 2 weeks. Logistic regression was used to analyze associations with outcomes of interest.

**Results:** Overall, 316 (10%) of 3,058 respondents wanted/needed a test in the two weeks prior to the survey. Of these, 166 (53%) were able to get tested and 156 (94%) received results; 53% waited ≥ 8 days to get results from when they wanted/needed a test. There were no significant differences by state. Among those wanting/needing a test, getting tested was significantly less common among men (aOR: 0.46) and those reporting black race (aOR: 0.53) and more common in those reporting recent travel (aOR: 3.35).

**Conclusions:** There is an urgent need for a national communication strategy on who should get tested and where one can get tested. Additionally, measures need to be taken to improve access and reduce turn-around-time.

## Introduction

While there is clear evidence supporting the need for rapid detection and isolation of SARS-CoV-2 infections,^1^ testing access across the US has been uneven,^2,3^ hampered by logistics, supply chain issues, and changing recommendations.^4^ For example, on August 24, 2020, the Centers for Disease Control and Prevention (CDC) recommended that asymptomatic persons not be tested even in the setting of potential exposure but reversed their recommendation on September 18, 2020 to reinforce testing for asymptomatic persons.^5^ In this dynamic context, community-based data on testing uptake and barriers are critical to the public health response. We present the SARS-CoV-2 testing cascade and associated barriers across three US states (Florida, Illinois, and Maryland) at varying local epidemic stages in September 2020.

## Methods

### Study Setting

We recruited participants from Florida, Illinois, and Maryland (∼1000/state) for an online survey

from September 16 – October 15, 2020. The survey covered demographics, COVID-19 symptoms, and experiences around SARS-CoV-2 PCR testing in the prior 2 weeks. States were selected to represent the diversity of the pandemic with respect to daily case counts and statewide orders on non-pharmaceutical interventions. At the time of survey, there were no systematic differences in testing across these states^5-7^; all had public and private options including free testing that did not require a doctor’s order. Cases counts were stable in Florida (∼2,700 cases/day), rising in Illinois (∼2,400 cases/day), and decreasing in Maryland (∼550 cases). Florida had the fewest restrictions (no restrictions on businesses or a statewide mask mandate), Maryland was fairly open with some restrictions (establishments up to 75% capacity, statewide mask mandate), and Illinois had many establishments open at 50% capacity, indoor dining at 25% capacity, and a statewide mask mandate.

### Study Sample

All participants were ≥ 18 years, provided consent, and resided in the state. Participants were recruited using Dynata (https://www.dynata.com), one of the largest first-party global data platforms. Dynata maintains a database of potential participants who are randomized to specific surveys if they meet the demographic targets of the survey; additionally, participants can select a survey from a list of potential options (survey topic not provided). Participants receive modest compensation. Security checks and quality verifications include digital fingerprinting and spot checking via third party verification.

In order to accrue demographically representative samples, we provided quotas for age, gender, race/ethnicity, and income based on the population composition of the states. Across states, 5,075 were routed to the survey; 714 did not start the survey, 694 started but did not complete the survey, and 609 responses were excluded for non-eligibility.

### Statistical Methods

Statistical analyses were carried out using Python (v3.7.3) and R (v3.5.1). We restricted the analyses to SARS-CoV-2 PCR testing in the prior two weeks to minimize recall bias and reflect current diagnostic testing access. Residential zip code and the National Center for Health Statistics (NCHS) Urban-Rural classification scheme^8^ were used for urban-rural classification. Chi square and Mann-Whitney tests were used to compare categorical and continuous variables, respectively. Logistic regression was used to analyze associations with two outcomes: 1) self-report of wanting/needing a SARS-CoV-2 test among all sampled; and 2) receiving a test among those who wanted/needed a test. Variables were considered for inclusion in multivariable analyses if they held biologic/epidemiologic significance or were associated in univariable analysis at p<0.10; age, gender, race/ethnicity, and state were included regardless of statistical significance. Additional variables considered included: household size, COVID-19 infection in a household member, education, annual household income, employment status (i.e., working outside the home), self-reported exposure and/or symptoms, report of travel, urban-rural classification, and state.

## Results

Of 3,058 persons, 316 (10%) reported wanting/needing a test in the prior two weeks. Median age of participants wanting/needing a test was 36 years and 46% were female; 47% self-identified as White and 57% reported working outside the home (overall sample described in Supplementary Table S1; Figure S2). In multivariable analyses, wanting/needing a test was significantly more common among males and younger respondents, Blacks vs. Whites, and those with symptoms, self-reported exposure or both (all p-values<0.05; Supplementary Table S3).

Of 316 who wanted/needed a test in the prior 2 weeks, 166 (53%) were able to get tested, of whom, 156 (94%) received results with no significant differences by state (Supplementary Figure S4). In multivariate analyses, among those wanting/needing a test, getting tested was significantly less common among men (aOR: 0.46; 95% CI: 0.26 – 0.82) and those reporting black race (aOR: 0.53; 95% CI: 0.28 – 0.99) and more common in those reporting recent travel (aOR: 3.35; 95% CI: 1.79 – 6.25) (Supplementary Table S5).

The primary reasons for testing were desire to know one’s status (35%) and symptoms (28%). Among those tested, 53% had to wait ≥ 8 days to get a result from when they wanted/needed a test (Table 1). Seventy-one percent reported quarantining while awaiting results. Of 146 who wanted/needed a test, but did not get tested, main reasons were not knowing where to go (36%) and distance/waiting time (33%); 21% reported fear of being tested.

**Table 1.**
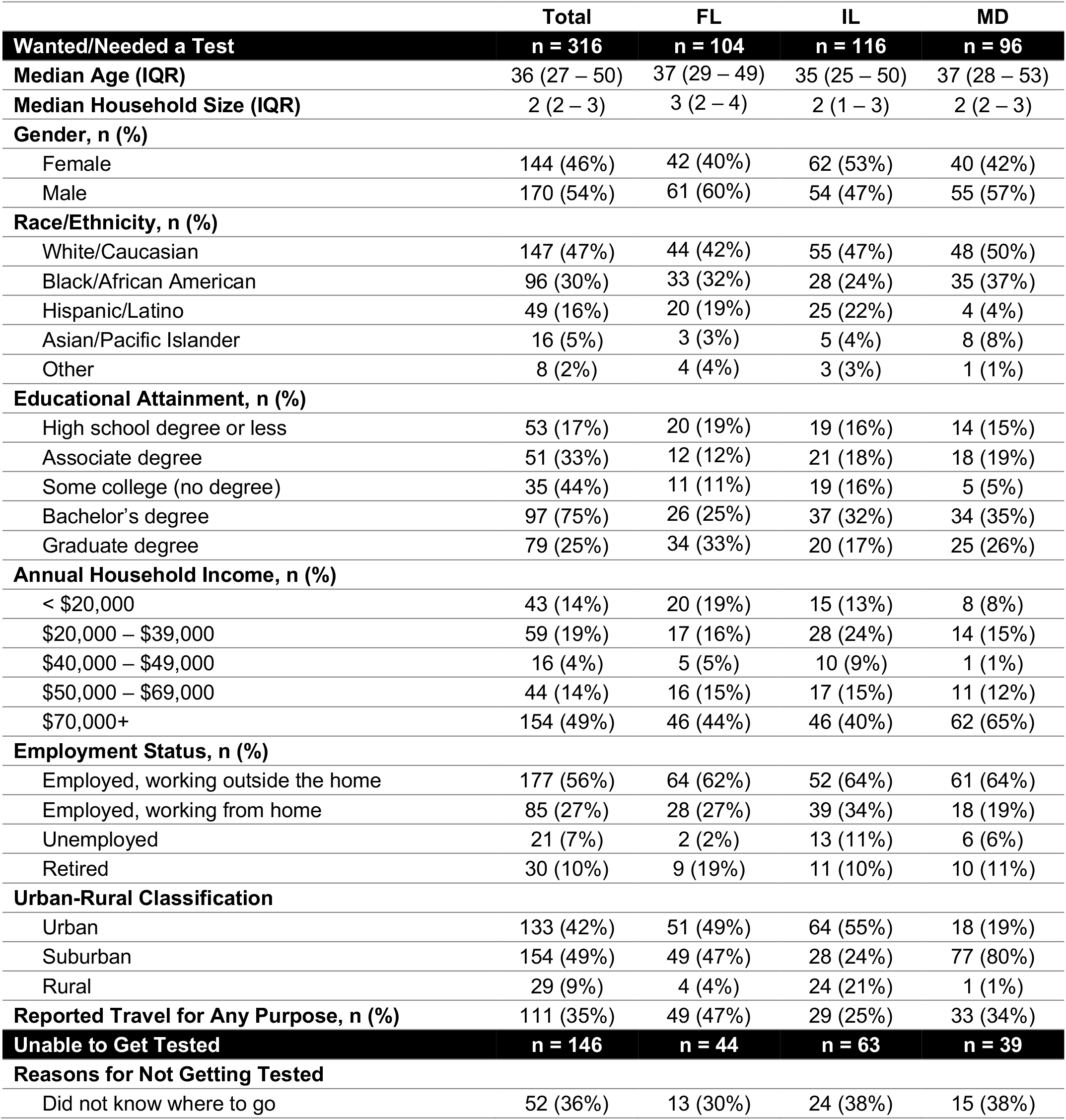
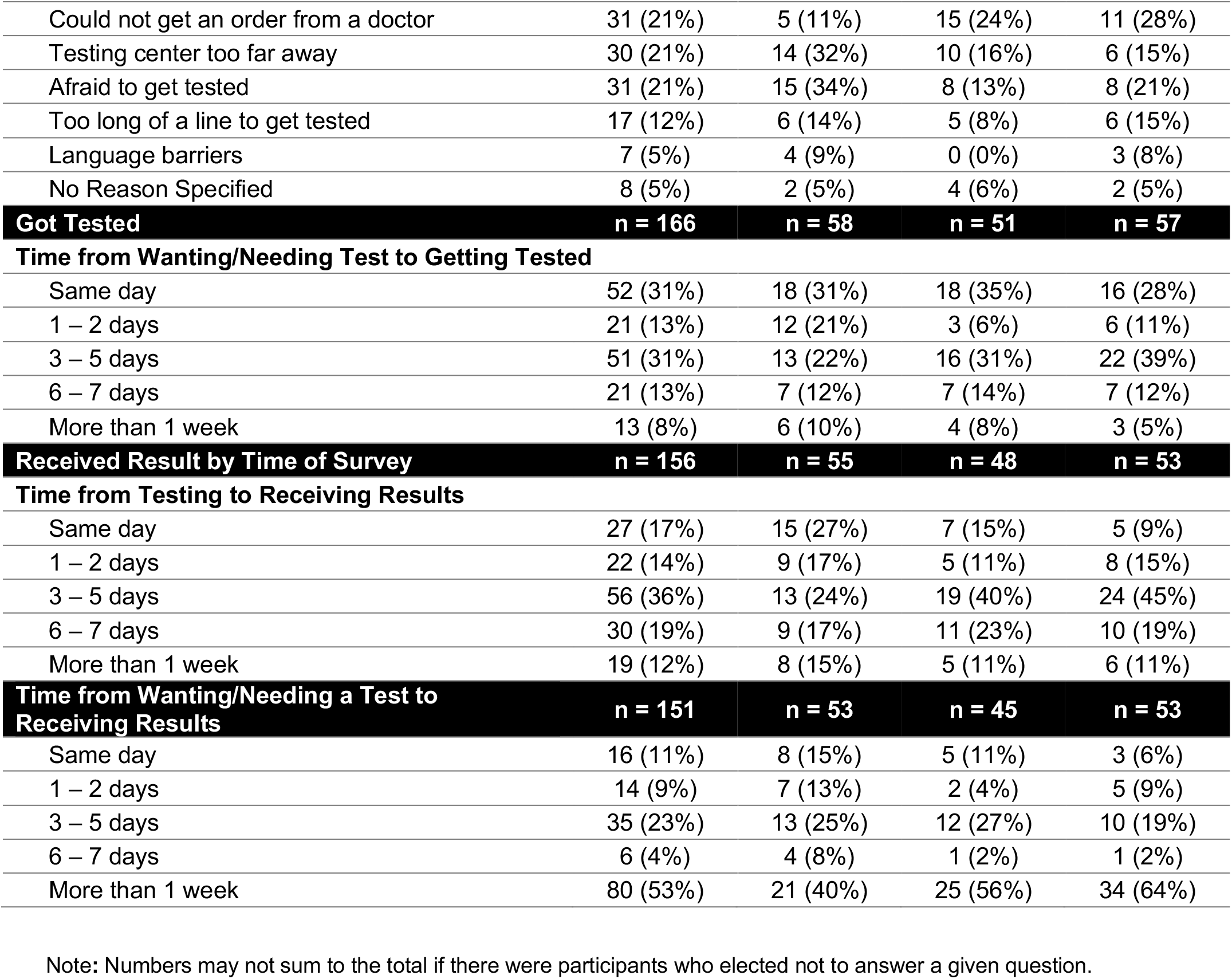
Demographics, barriers to testing, and wait times among persons who reported wanting/needing a SARS-CoV-2 PCR test in the prior 2 weeks across FL, IL, and MD.

Additionally, 177 (6%) participants reported symptoms, exposure or both but did not want a test. The main reasons were belief that symptoms were due to other causes (42%), no symptoms (18%), not wanting to know one’s status (18%), and logistic issues, such as not knowing where to go or lack of transportation (15%).

## Discussion

Of 3,058 participants surveyed, 10% reported wanting/needing a SARS-CoV-2 test in the prior two weeks, only 53% of who were able to get tested with no differences by state. While it was encouraging that those with symptoms and potential exposure were the most likely to get tested, it is noteworthy that approximately 62% with potential exposure and/or symptoms did not get tested primarily due to logistic reasons or not knowing where to go. Moreover, among those tested, there were significant delays in accessing a test and receiving results, with over half waiting 8 or more days for results from the time they wanted/needed a test. This delay is considerable compared to estimates of the serial interval (7.5 days)^9^ and generation time (5 days)^10^ which have implications for community transmission. Such delays can also complicate efforts to implement contact tracing to control local epidemics and may become more pronounced as cases again surge. Onset of respiratory viruses (e.g., influenza) can further complicate existing diagnostic efforts due to similar presenting symptoms thereby, increasing demand for testing.

There are limitations of online surveys; individuals need internet access to participate and so these surveys may underrepresent lower income/less educated individuals. However, in a constantly evolving pandemic where face-to-face data collection is nearly impossible, this approach allows for the collection of individual-level data across diverse geographies and demographics in a rapid and safe manner. If anything, we are likely overestimating access to testing. Moreover, care was taken to balance targets on state demographic characteristics and estimates of flu vaccine coverage are comparable to samples based on random digit dialing.^11^ Additionally, there is a possibility respondent misclassified the type of test.

Regardless, these data reflecting common testing barriers across three US states clearly underscore the importance of a unified national strategy with clear messaging on who, where, when, and how to get a test. Concurrently, there should be a focus on improved turn-around-time by incorporating newer approaches such as rapid lateral flow assays.

## Data Availability

Data are available from the corresponding author upon reasonable request.

## Acknowledgements

We would like to gratefully acknowledge Mr. Adebola Adegbesan who worked closely with our team in the recruitment of the study sample.

## Author Contributions

SJC, SSS, and SHM had full access to all the data in the study and take responsibility for the integrity of the data and the accuracy of the data analysis. *Concept and design*: SJC, AW, SHM, and SSS. *Acquisition, analysis, or interpretation of data*: All authors. *Drafting of the manuscript*: SJC, AW, SHM, SSS. *Critical revision of the manuscript for important intellectual content*: All authors. *Statistical analysis*: SJC. *Obtained funding:* SHM, AW. *Supervision:* SSS.

## Supplemental Material

**Table S1.** Characteristics of overall study sample by state.

|  | **Total** | **FL** | **IL** | **MD** |
| --- | --- | --- | --- | --- |
|  | **n = 3,058** | **n = 998** | **n = 1,045** | **n = 1,015** |
| **Median Age (IQR)** | 47 (33 – 64) | 48 (33 – 65) | 47 (35 – 64) | 46 (32 – 63) |
| **Median Household Size (IQR)** | 2 (2 – 3) | 2 (2 – 3) | 2 (1 – 3) | 2 (2 – 3) |
| **Gender, n (%)** |  |  |  |  |
| Female | 1,612 (53%) | 537 (51%) | 545 (52%) | 477 (53%) |
| Male | 1,431 (47%) | 505 (49%) | 502 (48%) | 422 (47%) |
| Other | 1 (0.03%) | 0 (0%) | 0 (0%) | 1 (0.1%) |
| **Race/Ethnicity, n (%)** |  |  |  |  |
| White/Caucasian | 1,712 (56%) | 444 (45%) | 628 (61%) | 640 (64%) |
| Black/African American | 632 (21%) | 223 (22%) | 169 (16%) | 240 (24%) |
| Hispanic/Latino | 437 (14%) | 241 (24%) | 141 (14%) | 55 (5%) |
| Asian/Pacific Islander | 175 (6%) | 58 (6%) | 65 (6%) | 52 (5%) |
| Other | 75 (3%) | 25 (3%) | 33 (3%) | 17 (2%) |
| **Educational Attainment, n (%)** |  |  |  |  |
| High school degree or less | 552 (18%) | 181 (18%) | 192 (18%) | 179 (18%) |
| Associate degree | 544 (18%) | 167 (17%) | 207 (20%) | 170 (17%) |
| Some college (no degree) | 335 (11%) | 133 (13%) | 122 (12%) | 80 (8%) |
| Bachelor's degree | 935 (31%) | 299 (30%) | 320 (31%) | 316 (31%) |
| Graduate degree | 672 (22%) | 213 (22%) | 198 (19%) | 261 (26%) |
| **Annual Household Income, n (%)** |  |  |  |  |
| < \$20,000 | 336 (11%) | 134 (13%) | 101 (10%) | 101 (10%) |
| \$20,000 – \$39,000 | 481 (16%) | 194 (19%) | 169 (16%) | 118 (12%) |
| \$40,000 – \$49,000 | 250 (8%) | 103 (10%) | 82 (8%) | 65 (6%) |
| \$50,000 – \$69,000 | 548 (18%) | 171 (17%) | 215 (21%) | 162 (16%) |
| \$70,000+ | 1,443 (47%) | 396 (40%) | 478 (46%) | 569 (56%) |
| **Employment Status, n (%)** |  |  |  |  |
| Employed, working outside the home | 1,168 (37%) | 361 (37%) | 427 (41%) | 380 (38%) |
| Employed, working from home | 878 (29%) | 275 (28%) | 299 (29%) | 304 (30%) |
| Unemployed | 309 (10%) | 102 (10%) | 110 (11%) | 97 (10%) |
| Retired | 664 (22%) | 245 (25%) | 199 (19%) | 220 (22%) |
| **Urban-Rural Classification** |  |  |  |  |
| Urban | 1,018 (33%) | 433 (43%) | 486 (47%) | 99 (10%) |
| Suburban | 1,764 (58%) | 511 (51%) | 385 (37%) | 868 (86%) |
| Rural | 276 (9%) | 54 (5%) | 174 (17%) | 48 (5%) |
| **Reported Exposure, n (%)** | 235 (8%) | 98 (10%) | 79 (8%) | 58 (6%) |
| **Reported Symptoms, n (%)** | 223 (7%) | 92 (9%) | 71 (7%) | 60 (6%) |
| **Reported Travel for Any Purpose, n (%)** | 494 (16%) | 184 (18%) | 142 (14%) | 168 (17%) |

Note: Numbers may not sum to the total if there were participants who elected not to answer a given question.

**Fig S2.**
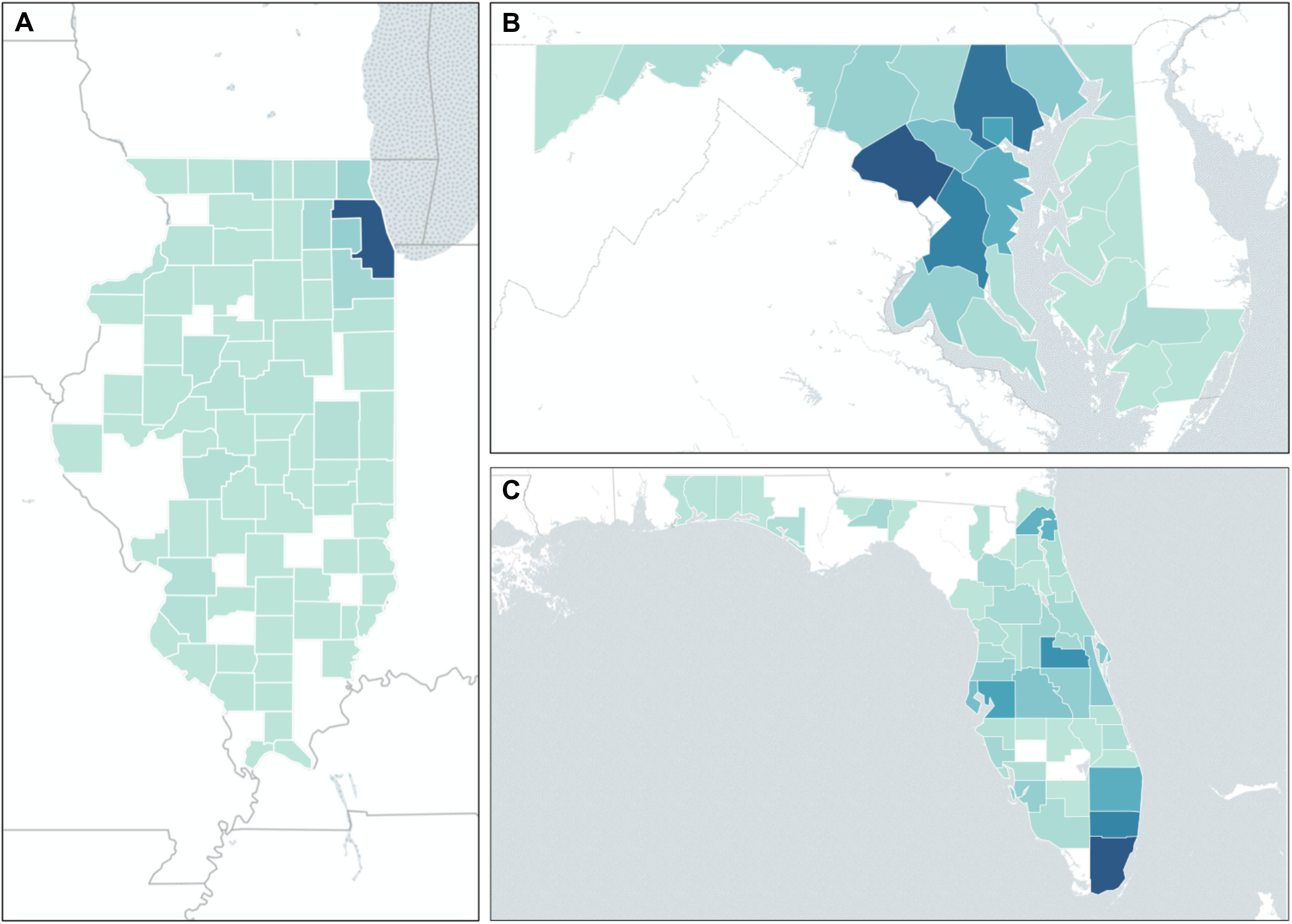
Distribution of study sample by county in each of the four study states: (A) Illinois, (B) Maryland, and (C) Florida. Darker shaded polygons represent a higher proportion of respondents.

**Table S3.** Factors associated with wanting/needing a SARS-CoV-2 PCR test in the prior 2 weeks (n=3,058). Results from univariable and multivariable regression analysis.

| Variable | Unadjusted Odds Ratio | 95% Confidence Interval | Adjusted Odds Ratio | 95% Confidence Interval |
| --- | --- | --- | --- | --- |
| **Age** (per 5-year increase) | 0.84 | 0.81 – 0.87 | 0.98 | 0.88 – 0.98 |
| **Household Size** (per 1-person increase) | 1.10 | 1.01 – 1.19 | - | - |
| **Positive Household Member**<br>(in the prior 2 weeks) | 11.04 | 7.93 – 15.4 | - | - |
| **Gender** |  |  |  |  |
| Female (ref.) | - | - | - | - |
| Male | 1.37 | 1.09 – 1.74 | 1.52 | 1.14 – 2.04 |
| **Race/Ethnicity** |  |  |  |  |
| White (ref.) | - | - | - | - |
| Black/African American | 1.93 | 1.47 – 2.54 | 1.85 | 1.29 – 2.67 |
| Hispanic/Latino | 1.35 | 0.96 – 1.91 | 1.34 | 0.86 – 2.07 |
| Asian/Pacific Islander | 1.07 | 0.62 – 1.84 | 1.30 | 0.70 – 2.42 |
| Other | 1.32 | 0.62 – 2.81 | 1.61 | 0.68 – 3.79 |
| **Educational Attainment** |  |  |  |  |
| High school degree or less (ref.) | - | - | - | - |
| Associate degree | 0.97 | 0.65 – 1.46 | - | - |
| Some college (no degree) | 1.09 | 0.70 – 1.72 | - | - |
| Bachelor's degree | 1.08 | 0.76 – 1.54 | - | - |
| Graduate degree | 1.24 | 0.86 – 1.79 | - | - |
| **Annual Household Income** |  |  |  |  |
| < \$20,000 (ref.) | - | - | - | - |
| \$20,000 – \$39,000 | 0.93 | 0.61 – 1.41 | - | - |
| \$40,000 – \$49,000 | 0.45 | 0.25 – 0.83 | - | - |
| \$50,000 – \$69,000 | 0.58 | 0.37 – 0.90 | - | - |
| \$70,000+ | 0.80 | 0.56 – 1.15 | - | - |
| **Employment Status** |  |  |  |  |
| Working Outside the Home (ref.) | - | - | - | - |
| Working from Home | 0.60 | 0.46 – 0.79 | 0.91 | 0.65 – 1.27 |
| Unemployed | 0.41 | 0.26 – 0.66 | 0.53 | 0.30 – 0.92 |
| Retired | 0.26 | 0.18 – 0.39 | 0.91 | 0.53 – 1.56 |
| **Urban-Rural Classification** |  |  |  |  |
| Urban (ref.) | - | - | - | - |
| Suburban | 0.63 | 0.49 – 0.81 | 0.79 | 0.57 – 1.10 |
| Rural | 0.77 | 0.50 – 1.18 | 0.79 | 0.45 – 1.37 |
| **State** |  |  |  |  |
| FL (ref.) | - | - | - | - |
| IL | 1.07 | 0.81 – 1.41 | 1.49 | 1.04 – 2.14 |
| MD | 0.89 | 0.67 – 1.20 | 1.40 | 0.95 – 2.06 |
| **Report Exposure and/or Symptoms** |  |  |  |  |
| No exposure or symptoms (ref.) | - | - | - | - |
| Exposure only | 10.5 | 7.07 – 15.6 | 8.27 | 5.45 – 12.6 |
| Symptoms only | 12.2 | 8.10 – 18.4 | 10.1 | 6.48 – 15.7 |
| Exposure and symptoms | 64.2 | 38.7 – 106 | 53.2 | 30.7 – 92.1 |
| **Report Travel for Any Purpose** | 3.29 | 2.55 – 4.24 | 1.34 | 0.95 – 1.90 |

**Figure S4.**
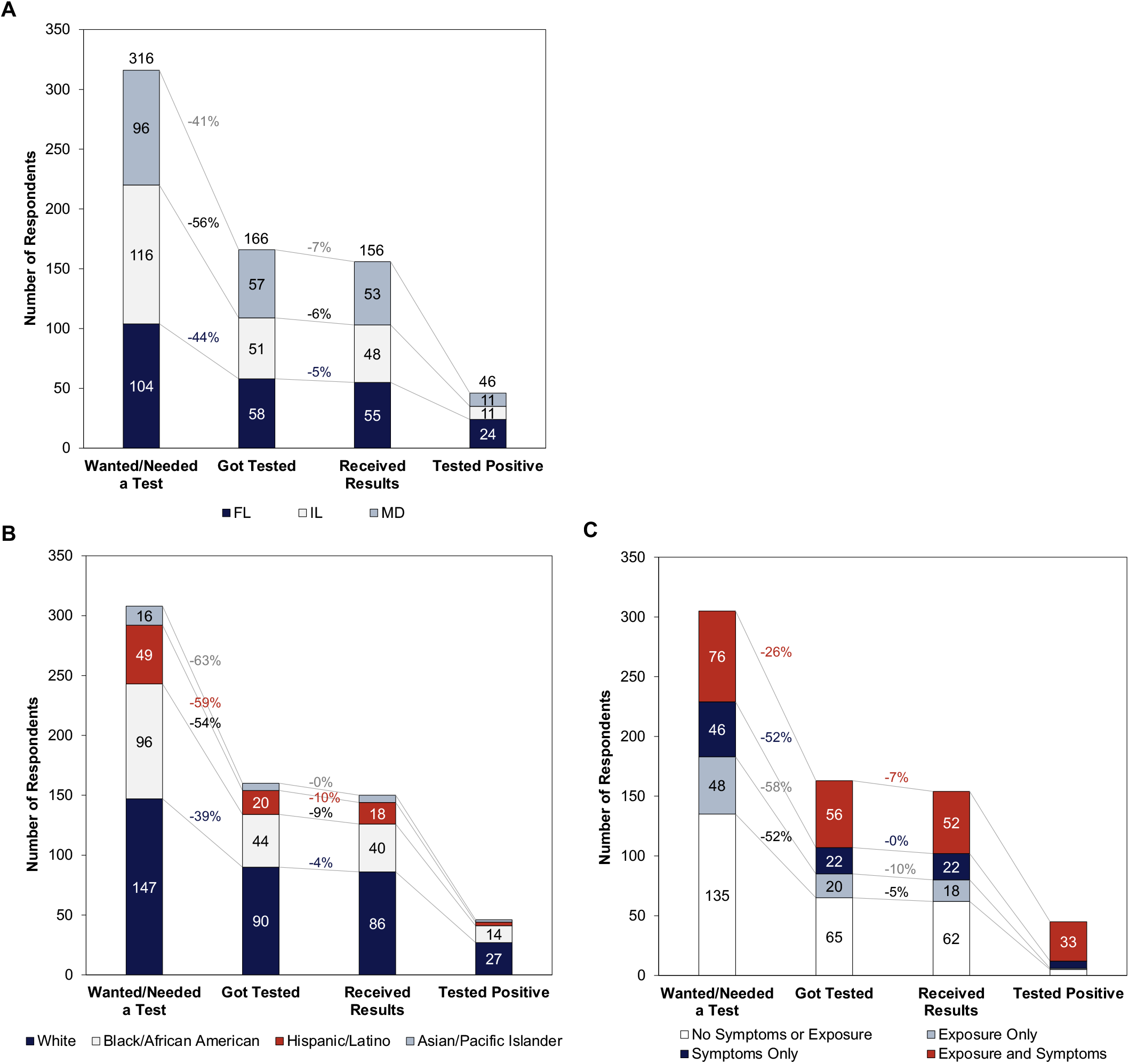
SARS-CoV-2 testing cascade by (A) state, (B) race/ethnicity, and (C) self-reported symptoms. Trend lines and percentages reflect the proportion lost between each step in the cascade. Note: Numbers may not sum to the total if there were participants who elected not to answer a given question.

**Table S5.** Factors associated with getting a PCR test for SARS-CoV-2 among those who wanted/needed a test in the prior 2 weeks (n=316). Results from univariable and multivariable logistic regression analysis.

| Variable | Unadjusted Odds Ratio | 95% Confidence Interval | Adjusted Odds Ratio | 95% Confidence Interval |
| --- | --- | --- | --- | --- |
| **Age** (per 5-year increase) | 1.00 | 0.94 – 1.08 | 1.00 | 0.90 – 1.10 |
| **Household Size** (per 1-person increase) | 0.99 | 0.85 – 1.14 | - | - |
| **Positive Household Member**<br>(in the prior 2 weeks) | 3.13 | 1.79 – 5.50 | - | - |
| **Gender** |  |  |  |  |
| Female (ref.) | - | - | - | - |
| Male | 0.93 | 0.59 – 1.45 | 0.46 | 0.26 – 0.82 |
| **Race/Ethnicity** |  |  |  |  |
| White (ref.) | - | - | - | - |
| Black/African American | 0.56 | 0.33 – 0.94 | 0.53 | 0.28 – 0.99 |
| Hispanic/Latino | 0.45 | 0.23 – 0.88 | 0.60 | 0.28 – 1.28 |
| Asian/Pacific Islander | 0.42 | 0.14 – 1.25 | 0.34 | 0.10 – 1.13 |
| Other | 1.90 | 0.37 – 9.74 | 2.37 | 0.35 – 16.2 |
| **Educational Attainment** |  |  |  |  |
| High school degree or less (ref.) | - | - | - | - |
| Associate degree | 0.89 | 0.56 – 1.41 | - | - |
| Some college (no degree) | 1.11 | 0.67 – 1.83 | - | - |
| Bachelor's degree | 1.36 | 0.93 – 2.00 | - | - |
| Graduate degree | 1.58 | 1.06 – 2.34 | - | - |
| **Annual Household Income** |  |  |  |  |
| < \$20,000 (ref.) | - | - | - | - |
| \$20,000 – \$39,000 | 0.70 | 0.32 – 1.56 | - | - |
| \$40,000 – \$49,000 | 0.66 | 0.20 – 2.15 | - | - |
| \$50,000 – \$69,000 | 0.43 | 0.18 – 1.02 | - | - |
| \$70,000+ | 1.10 | 0.55 – 2.20 | - | - |
| **Employment Status** |  |  |  |  |
| Working Outside the Home (ref.) | - | - | - | - |
| Working from Home | 0.51 | 0.30 – 0.86 | 1.54 | 0.83 – 2.85 |
| Unemployed | 0.25 | 0.09 – 0.68 | 0.36 | 0.11 – 1.22 |
| Retired | 0.55 | 0.25 – 1.20 | 1.27 | 0.43 – 3.69 |
| **Urban-Rural Classification** |  |  |  |  |
| Urban (ref.) | - | - | - | - |
| Suburban | 1.22 | 0.76 – 1.95 | 0.68 | 0.37 – 1.23 |
| Rural | 1.42 | 0.63 – 3.20 | 1.72 | 0.64 – 4.60 |
| **State** |  |  |  |  |
| FL (ref.) | - | - | - | - |
| IL | 0.61 | 0.36 – 1.05 | 0.75 | 0.38 – 1.46 |
| MD | 1.11 | 0.63 – 1.95 | 1.64 | 0.82 – 3.30 |
| **Report Exposure and/or Symptoms** |  |  |  |  |
| No exposure or symptoms (ref.) | - | - | - | - |
| Exposure only | 0.75 | 0.38 – 1.46 | 0.66 | 0.31 – 1.38 |
| Symptoms only | 0.96 | 0.49 – 1.88 | 0.70 | 0.32 – 1.49 |
| Exposure and symptoms | 3.08 | 1.66 – 5.74 | 1.84 | 0.90 – 3.79 |
| **Report Travel for Any Purpose** | 3.66 | 2.20 – 6.09 | 3.35 | 1.79 – 6.25 |

**Table S6.** Barriers to testing by symptoms among persons who reported wanting/needing a SARS-CoV-2 PCR test in the prior 2 weeks across FL, IL, and MD.

|  | Total | No Symptoms | Any Symptoms |
| --- | --- | --- | --- |
| Unable to Get Tested | n = 146 | n = 99 | n = 45 |
| **Reasons for Not Getting Tested** |  |  |  |
| Did not know where to go | 52 (36%) | 31 (31%) | 20 (44%) |
| Too long of a line to get tested | 17 (12%) | 12 (112%) | 5 (11%) |
| Could not get an order from a doctor | 31 (21%) | 19 (19%) | 12 (27%) |
| Testing center too far away | 31 (21%) | 24 (24%) | 7 (16%) |
| Afraid to get tested | 30 (21%) | 17 (17%) | 13 (29%) |
| Language barriers | 7 (5%) | 3 (3%) | 4 (9%) |

